# Impact of diabetes mellitus on the stabilization and osseointegration of dental implants: a systematic review

**DOI:** 10.1101/2022.02.28.22271622

**Authors:** Aglaia Katsiroumpa, Petros Galanis, Ioannis Diamantis, Sofia Georgikopoulou, Theodoros Katsoulas, Elissavet Katsimperi, Evangelos Konstantinou

## Abstract

**Introduction:** Diabetes mellitus is a chronic metabolic disorder leading to hyperglycemia, which causes various complications due to vasculopathy. Adequate dental rehabilitation including dental implants plays a key role in promoting the eating habits of diabetics and better metabolic control.

**Aim:** We performed a systematic literature review to investigate the effect of diabetes mellitus on the stabilization and osseointegration of dental implants.

**Methods:** We searched PubMed and Scopus until October 2021. We used the following inclusion criteria: (i) study population included diabetics type I or II, (ii) outcomes were the dental implant failure or resonance frequency analysis, (iii) studies that investigate the effect of diabetes mellitus on the stabilization and osseointegration of dental implants, (iv) studies that were published in English, (v) studies that were published in journals with peer review system, (vi) studies including humans and not animals, (vii) studies that compared diabetics with non-diabetics, and (ix) quantitative studies.

**Results:** Twenty-nine studies met the inclusion criteria. Regarding implant failure, four studies found statistically significant more frequent implant failure in diabetics, while five studies found that implant failure was more frequent in diabetics but was not statistically significant. In contrast, ten studies found that implant failure was more frequent in non-diabetics but was not statistically significant. In addition, seven studies found that all diabetics and non-diabetics retained their implant during the study. In six studies that performed the resonance frequency analysis, no statistically significant difference was found between diabetics and non-diabetics. In three studies, the mean value of the implant stability quotient increased statistically significant in non-diabetics, while in three studies the mean value of the implant stability quotient increased statistically significant in diabetics.

**Conclusions:** The results of this review suggest that implant failure is not higher for diabetics than for non-diabetics. Diabetics seem to be able to achieve a rate of dental implants survival like that of non-diabetics. With regards to the resonance frequency analysis, no difference is found between diabetics and non-diabetics.

## Introduction

Dental implants are a method to restore lost teeth. Advances in dental research and implant creation have established implants as a highly effective method. In particular, the average implant survival rate reaches 94.6% even after 10 years of implant placement (Moraschini et al., 2015). The survival of an implant initially depends on its successful osseointegration after its placement. Several factors influence implant survival with diabetes mellitus being a potential factor to be investigated.

Diabetes mellitus is a chronic metabolic disorder leading to hyperglycemia, which causes various complications due to vasculopathy. Diabetics have an increased incidence of periodontitis and tooth loss, delayed wound healing and worse outcomes in infections (Abiko & Selimovic, 2010; Khader et al., 2006). The prevalence of diabetes is continuously increasing. For example, in 1980, more than 150 million people worldwide had diabetes, and in 2008, this number exceeded 350 million people (Danaei et al., 2011). For this reason, a better understanding of diabetes and its treatment, as well as its impact on the outcome of dental implants, is essential.

The role of dental implants in diabetics is extremely important, as these patients, after tooth loss, avoid foods that cause them difficulty in chewing, resulting in an inappropriate diet. Adequate dental rehabilitation with the use of implants plays a key role in promoting the eating habits of diabetics and better metabolic control. Identifying the factors that increase the risk of complications in dental patients enables surgeons to make rational decisions according to the evidence and determine the best possible plan of care, achieving the best clinical outcomes (Chrcanovic et al., 2014).

The aim of this systematic literature review was to investigate the effect of diabetes mellitus on the stabilization and osseointegration of dental implants.

## Methods

We searched PubMed and Scopus until October 2021. We used the following inclusion criteria: (i) study population included diabetics type I or II, (ii) outcomes were the dental implant failure or resonance frequency analysis, (iii) studies that investigate the effect of diabetes mellitus on the stabilization and osseointegration of dental implants, (iv) studies that were published in English, (v) studies that were published in journals with peer review system, (vi) studies including humans and not animals, (vii) studies that compared diabetics with non-diabetics, and (ix) quantitative studies.

We applied the Preferred Reporting Items for Systematic Reviews and Meta-Analyses (PRISMA) guidelines. PICO methodology was used to create the search strategy (Table 1). We used the following search strategy: ((“dental implant” OR “dental implant surgery”) AND (diabetic* OR “diabetes mellitus” OR “type 1 diabetes mellitus” OR “type 2 diabetes mellitus” OR “diabetic type 1” OR “diabetic type 2”)) AND (“resonance frequency analysis” OR RFA OR survival OR “dental implant survival” OR failure OR “dental implant failure”).

**Table 1.** Search strategy in PubMed κα*l* Scopus using PICO methodology.

| **PICO** | **Keywords** |
| --- | --- |
| P | dental implant OR dental implant surgery |
| I | diabetic* OR diabetes mellitus OR type 1 diabetes mellitus OR type 2<br>diabetes mellitus OR diabetic type 1 OR diabetic type 2 |
| C | non-diabetic* |
| O | resonance frequency analysis OR RFA OR survival OR dental implant<br>survival OR failure OR dental implant failure |

Flowchart of the systematic literature review is presented in Figure 1. Initially, we found 68 records in PubMed and 2080 record in Scopus. Applying inclusion criteria, 29 studies included in our review.

**Figure 1.**
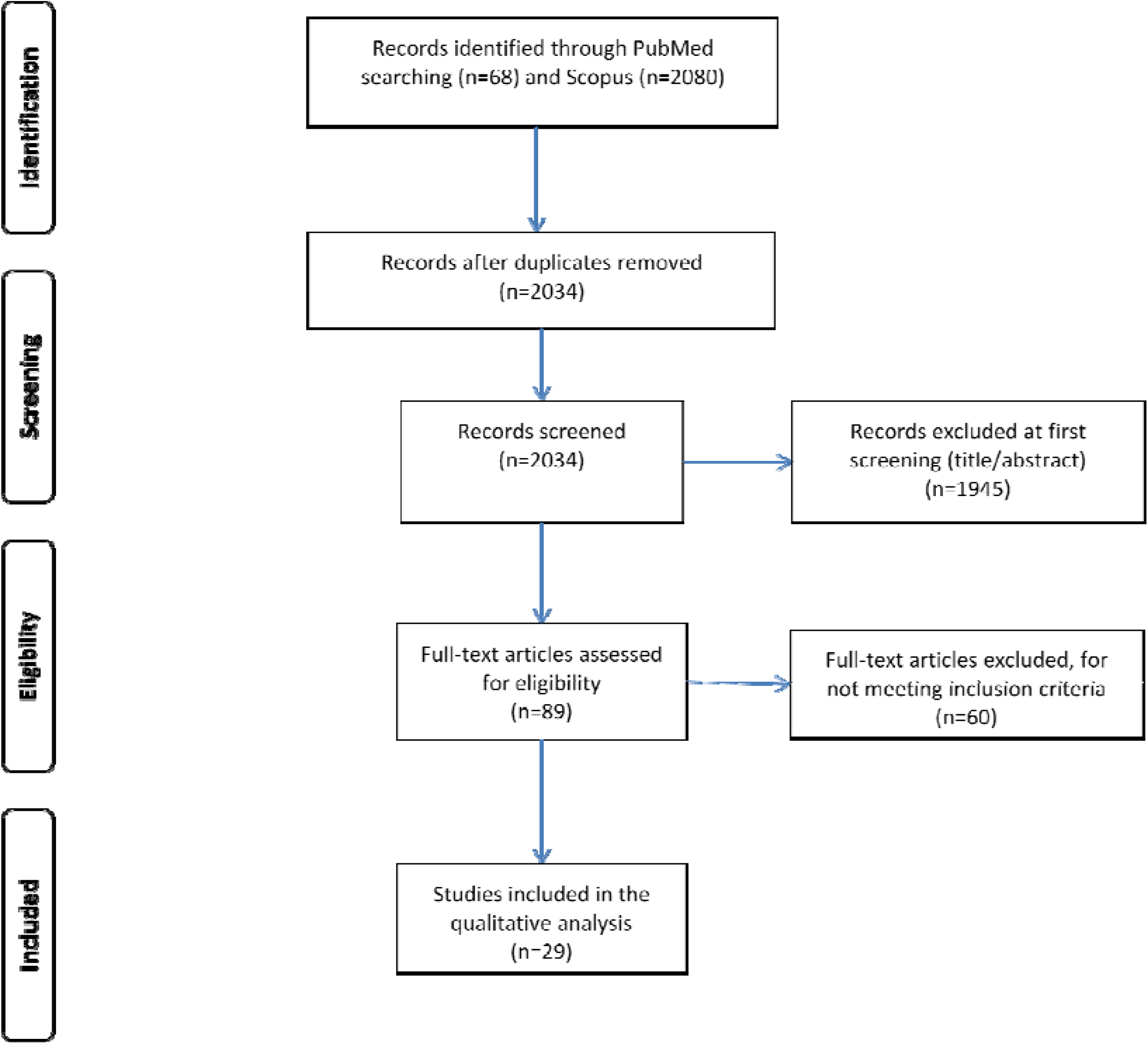
Flowchart of the systematic literature review.

## Results

Twenty-nine studies met the inclusion criteria. Detailed characteristics of the studies included in the systematic literature review are presented in Table 2. The majority of studies were conducted in Asia (n=12) and the USA (n=11), while five studies were conducted in Europe and one study in Brazil.

**Table 2.**
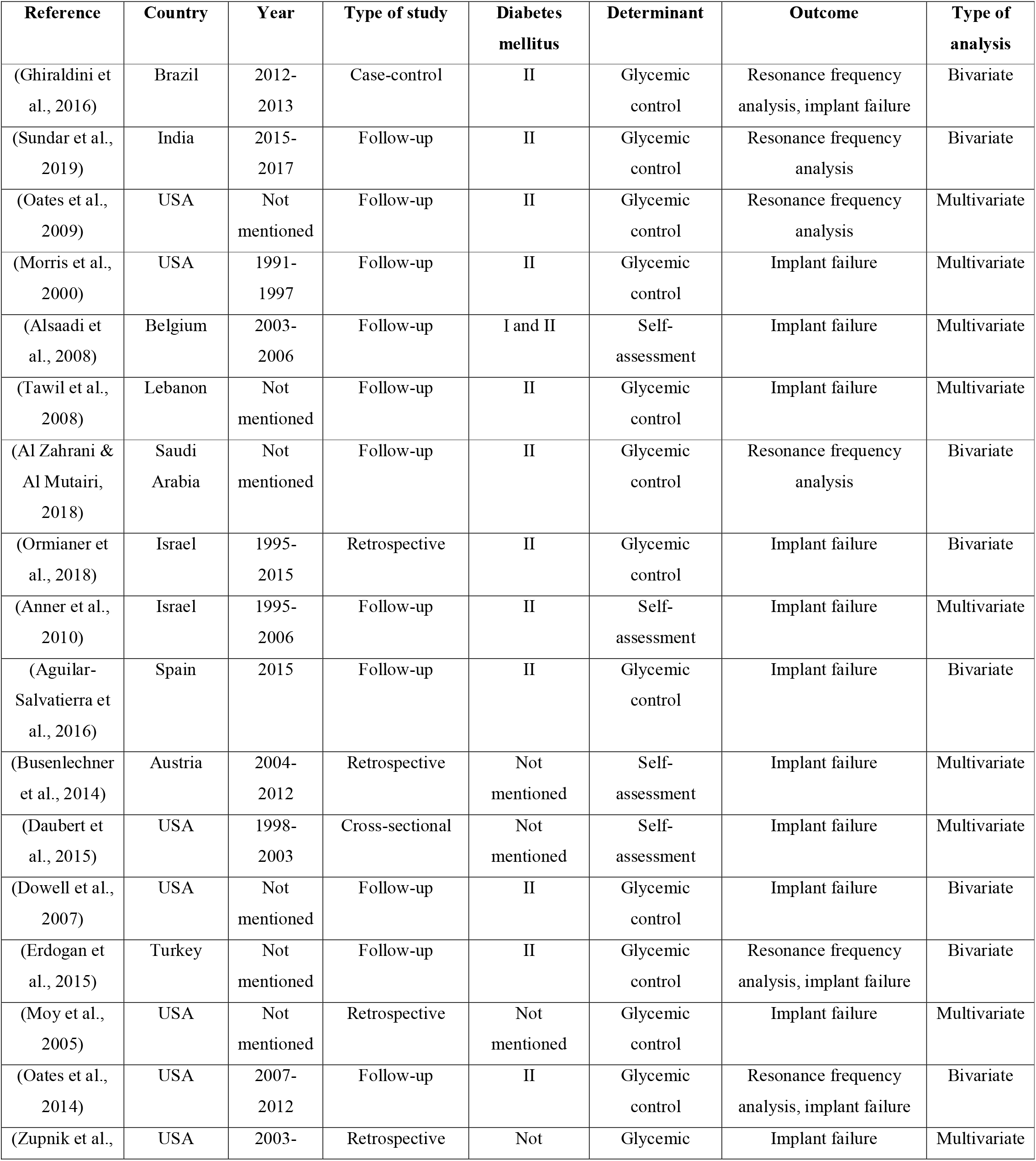
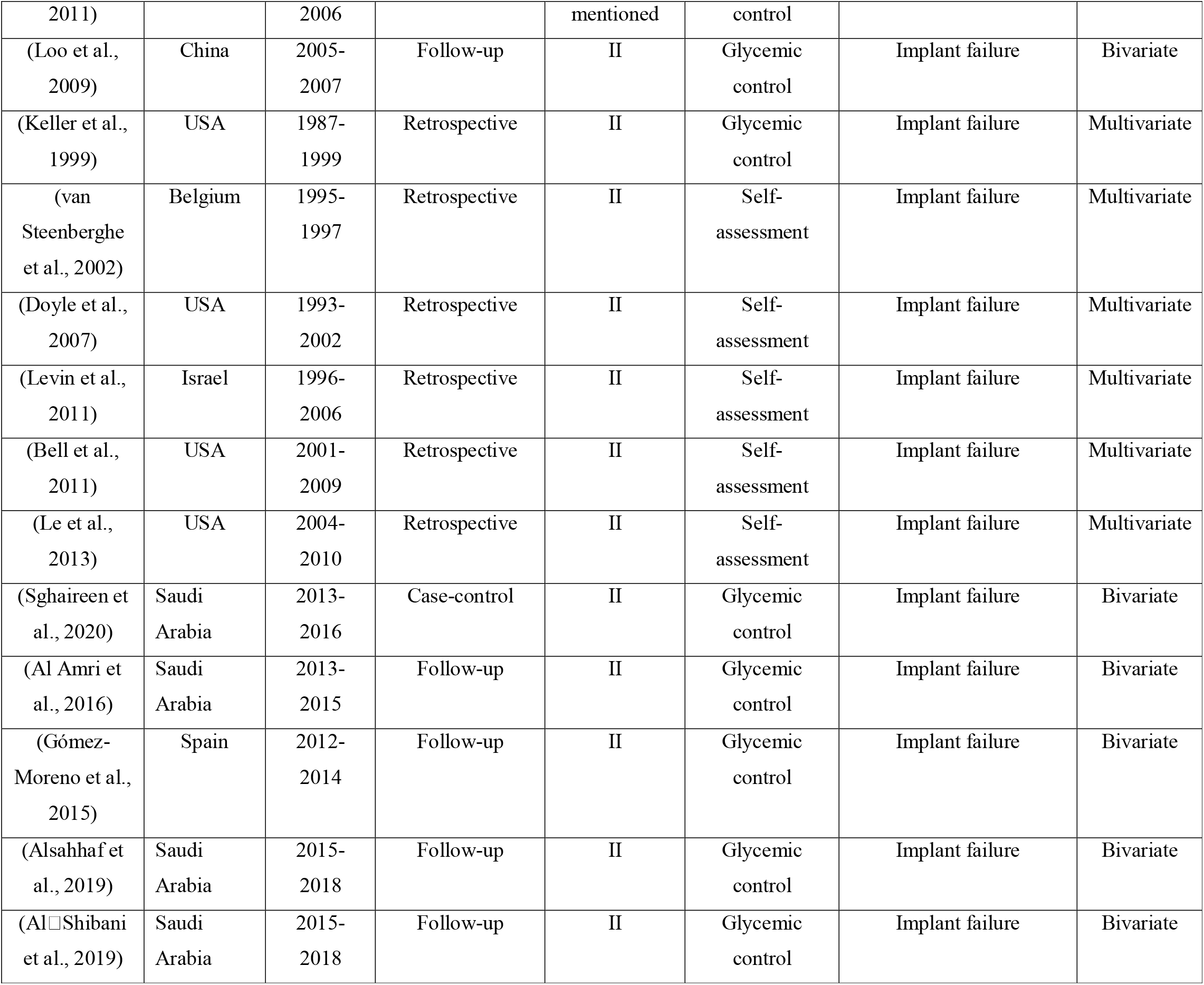
Detailed characteristics of the studies included in the systematic literature review.

Most of the studies were follow-up studies (n=16) and retrospective studies (n=10), while two studies were case-control, and one study was cross-sectional. Study population included mainly diabetics type II (n=24), while one study included diabetics type I and II. Four studies did not mention the type of diabetes mellitus. Glycemic control was used in 20 studies to clarify the diabetic status of the participants, while self-assessment was used in nine studies.

The outcome in most studies (n=23) was the dental implant failure, while the resonance frequency analysis was used in three studies. Also, three studies measured both the dental implant failure and the resonance frequency analysis. Multivariate analysis was used in 15 studies eliminating confounders, while bivariate analysis was used in 14 studies.

Detailed results of the studies included in the systematic literature review are shown in Table 3. Regarding implant failure, four studies found statistically significantly more frequent implant failure in diabetics (Daubert et al., 2015; Loo et al., 2009; Moy et al., 2005; Zupnik et al., 2011), while five studies found that implant failure was more frequent in diabetics but was not statistically significant (Aguilar-Salvatierra et al., 2016; Morris et al., 2000; Ormianer et al., 2018; Sghaireen et al., 2020; Tawil et al., 2008). In contrast, ten studies found that implant failure was more frequent in non-diabetics but was not statistically significant (Alsaadi et al., 2008; Anner et al., 2010; Bell et al., 2011; Busenlechner et al., 2014; Doyle et al., 2007; Keller et al., 1999; Le et al., 2013; Levin et al., 2011; Oates et al., 2014; van Steenberghe et al., 2002). In addition, seven studies found that all diabetics and non-diabetics retained their implant during the study (Al Amri et al., 2016; Alsahhaf et al., 2019; Al□Shibani et al., 2019; Dowell et al., 2007; Erdogan et al., 2015; Gómez-Moreno et al., 2015; Sundar et al., 2019). In six studies that performed the resonance frequency analysis, no statistically significant difference was found between diabetics and non-diabetics (Al Zahrani & Al Mutairi, 2018; Erdogan et al., 2015; Ghiraldini et al., 2016; Oates et al., 2009, 2014; Sundar et al., 2019). In three studies, the mean value of the implant stability quotient increased statistically significant in non-diabetics (Ghiraldini et al., 2016; Oates et al., 2014; Sundar et al., 2019), while in three studies the mean value of the implant stability quotient increased statistically significant in diabetics (Al Zahrani & Al Mutairi, 2018; Oates et al., 2014; Sundar et al., 2019).

**Table 3.** Detailed results of the studies included in the systematic literature review.

| **Reference** | **Statistically significant difference** | **Greater improvement in** | **Percentage of dental implant failure** |
| --- | --- | --- | --- |
| (Ghiraldini et al., 2016) | No | None | Not mentioned |
| (Sundar et al., 2019) | No | None | 0% in diabetics and non-diabetics |
| (Oates et al., 2009) | No | None | Not mentioned |
| (Morris et al., 2000) | No | Non-diabetics | Not mentioned |
| (Alsaadi et al., 2008) | No | Diabetics | Not mentioned |
| (Tawil et al., 2008) | No | Non-diabetics | Not mentioned |
| (Al Zahrani & Al Mutairi, 2018) | No | None | Not mentioned |
| (Ormianer et al., 2018) | No | Non-diabetics | 6% in diabetics and 4.4% in non-diabetics |
| (Anner et al., 2010) | No | Diabetics | Not mentioned |
| (Aguilar-Salvatierra et al., 2016) | No | Non-diabetics | 3.4% in diabetics and 0% in non-diabetics |
| (Busenlechner et al., 2014) | No | Diabetics | 3% in diabetics and 4.9% in non-diabetics |
| (Daubert et al., 2015) | Yes | Non-diabetics | Not mentioned |
| (Dowell et al., 2007) | No | None | 0% in diabetics and non-diabetics |
| (Erdogan et al., 2015) | No | None | 0% in diabetics and non-diabetics |
| (Moy et al., 2005) | Yes | Non-diabetics | Not mentioned |
| (Oates et al., 2014) | No | Non-diabetics | 0% in diabetics and 1% in non-diabetics |
| (Zupnik et al., 2011) | Yes | Non-diabetics | Not mentioned |
| (Loo et al., 2009) | Yes | Non-diabetics | Not mentioned |
| (Keller et al., 1999) | No | Diabetics | Not mentioned |
| (van Steenberghe et al., 2002) | No | Diabetics | Not mentioned |
| (Doyle et al., 2007) | No | Diabetics | Not mentioned |
| (Levin et al., 2011) | No | Diabetics | Not mentioned |
| (Bell et al., 2011) | No | Diabetics | Not mentioned |
| (Le et al., 2013) | No | Diabetics | Not mentioned |
| (Sghaireen et al., 2020) | No | Non-diabetics | Not mentioned |
| (Al Amri et al., 2016) | No | None | 0% in diabetics and non-diabetics |
| (Gómez-Moreno et al., 2015) | No | None | 0% in diabetics and non-diabetics |
| (Alsahhaf et al., 2019) | No | None | 0% in diabetics and non-diabetics |
| (Al-Shibani et al., 2019) | No | None | 0% in diabetics and non-diabetics |

## Discussion

We performed a systematic literature review to investigate the effect of diabetes mellitus on the stabilization and osseointegration of dental implants. In general, diabetes mellitus does not seem to affect the stabilization and osseointegration of dental implants since only four studies found that implant failure was statistically significant more frequent in diabetics. Also, five studies found that implant failure was more frequent in diabetics but was not statistically significant, but ten studies found that implant failure was more frequent in non-diabetics but was not statistically significant. Moreover, seven studies found that all diabetics and non-diabetics retained their implant during the study.

Other systematic reviews found similar findings with us (Andrade et al., 2021; Chrcanovic et al., 2014; Shang & Gao, 2021). In particular, Chrcanovic et al. (2014) found that the diabetic status does not significantly affect implant failure rates (odds ratio = 1.07, 95% confidence interval = 0.8 to 1.44, p-value = 0.65). On the other hand, Chrcanovic et al. (2014) found a statistically significant difference between diabetics and non-diabetics regarding marginal bone loss, in favor of non-diabetics (mean difference = 0.2, 95% confidence interval = 0.08 to 0.31, p-value = 0.001). Shang & Gao (2021) did not find significant differences in rates of implant failure (odds ratio = 1.39, 95% confidence interval = 0.58 to 3.3, p-value = 0.46) and probing death (mean difference = 0.2, 95% confidence interval = −0.04 to 0.44, p-value = 0.1) between diabetics and non-diabetics, but they found significant differences in peri-implant bleeding on probing (mean difference = 0.32, 95% confidence interval = 0.19 to 0.45, p-value < 0.001) and peri-implant bone loss (mean difference = 0.12, 95% confidence interval = 0.02 to 0.22, p-value = 0.02), favoring non-diabetics. Meta-analysis of Andrade et al. (2021) showed no significant difference between diabetics and non-diabetics regarding marginal bone loss (mean difference = −0.08, 95% confidence interval = −0.25 to 0.08, p-value = 0.33) and implant survival rates (odds ratio = 1.0, 95% confidence interval = 0.96 to 1.04, p-value = 0.91) even in diabetics with poor glycemic control (odds ratio = 1.08, 95% confidence interval = 0.87 to 1.33, p-value = 0.48).

There is a disagreement among studies about what occurs with the uncontrolled diabetic patients. Several studies have shown the unsatisfactory outcomes of dental treatment in diabetics with poor glycemic control (de Lima et al., 2020; Lagunov et al., 2019; Quirino et al., 1995), but a meta-analysis found that the dental implant survival rate was similar in uncontrolled diabetics and non-diabetics (Andrade et al., 2021). Moreover, uncontrolled diabetics have higher values of marginal bone loss, bleeding on probing, and pocket depth (Aguilar-Salvatierra et al., 2016; Al Amri et al., 2016). Satisfactory glycemic control in diabetic patients is essential since the HbA1c level is related with peri-impant pathology (Ibraheem et al., 2019; Javed & Romanos, 2009).

Diabetes considered being a contraindication for treatment with implants (Michaeli et al., 2009), but success rates among diabetics with controlled glucose may be similar to those of non-diabetics (Ciancio et al., 1995; Oates et al., 2013). However, a few studies only in this review monitored the glycemic control throughout the follow-up study. Moreover, in nine studies the measurement of glucose was not even carried out at the beginning of the study and the diabetic status was defined through self-assessment. This fact may have lead to confusion in our review. Satisfactory glycemic control is related with high implant survival rate probably due to the absence of bacteria and their products in systemic circulation (Al Amri et al., 2016; Javed & Romanos, 2009).

## Limitations

Our review had several limitations. Study population included diabetics type II in 24 studies, while one study included diabetics type I and II and four studies did not mention the type of diabetes mellitus. Therefore, more studies should be carried out with type 1 diabetics in order to draw safer conclusions about these patients. Moreover, an information bias could be introduced since the diabetic status of patients was defined through self-assessment in nine studies. The resonance frequency analysis to measure objectively the stabilization of dental implants was used only in six studies. Also, almost half of the studies in our review did not use multivariate analysis to eliminate confounders and only crude measures of effect were estimated. Possible confounding factors can influence the impact of diabetes mellitus on the stabilization and osseointegration of dental implants. Since the effect of diabetes mellitus on the stabilization and osseointegration of dental implants remains unclear, randomized controlled trials examining the influence of diabetes on the survival of dental implants should be conducted as soon as possible. Furthermore, possible confounding should be eliminated to minimize bias.

## Conclusions

The results of this systematic literature review suggest that implant failure is not higher for diabetics than for non-diabetics. Diabetics seem to be able to achieve a rate of dental implants survival like that of non-diabetics. With regards to the resonance frequency analysis, no difference is found between diabetics and non-diabetics. Moreover, in three studies, the mean value of the implant stability quotient increased statistically significant in non-diabetics, while three studies arrived at the exact opposite conclusion. A greater number of well-designed randomized controlled trials are required to draw safer conclusions.

## Data Availability

All data produced in the present study are available upon reasonable request to the authors

